# Estimating diagnostic noise in panel-based genomic analysis

**DOI:** 10.1101/2022.03.18.22272595

**Authors:** Robin N. Beaumont, Caroline F. Wright

## Abstract

**Background:** Gene panels with a series of strict variant filtering rules are often used for clinical analysis of exomes and genomes. Panels vary in size, which affects the sensitivity and specificity of the test. We sought to investigate the background rate of candidate diagnostic variants in a population setting using gene panels developed to diagnose a range of heterogeneous monogenic diseases.

**Methods:** We used the Genotype-2-Phenotype database with the Variant Effect Predictor plugin to identify rare non-synonymous variants in exome sequence data from 200,643 individuals in UK Biobank. We evaluated five clinically curated gene panels: developmental disorders (DD; 1708 genes), heritable eye disease (536 genes), skin disorders (293 genes), cancer syndromes (91 genes) and cardiac conditions (49 genes). We further tested the DD panel in 9,860 proband-parent trios from the Deciphering Developmental Disorders (DDD) study.

**Results:** As expected, bigger gene panels resulted in more variants being prioritised, varying from an average of ∼0.3 per person in the smallest panels, to ∼3.5 variants per person using the largest panel. The number of individuals with prioritised variants varied linearly with coding sequence length for monoallelic disease genes (∼300 individuals per 1000 base pairs) and quadratically for biallelic disease genes, with some notable outliers. Based on cancer registry data from UK Biobank, there was no detectable difference between cases and controls in the number of individuals with prioritised variants using the cancer panel, presumably due to the predominance of sporadic disease. However, we observed a marked increase in the number of prioritised variants in the DD panel in the DDD study (∼5 variants per proband). Phasing of compound heterozygotes in biallelic genes resulted in a modest reduction in the number of prioritised variants.

**Conclusions:** Although large gene panels may be the best strategy to maximize diagnostic yield in genetically heterogeneous diseases, they will frequently prioritise false positive candidate variants potentially requiring additional clinical follow-up. Most individuals will have at least one rare nonsynonymous variant in panels containing >500 monogenic disease genes. Extreme caution should therefore be applied when interpreting potentially pathogenic variants found in the absence of relevant phenotypes.

## Background

Clinical analysis of exomes and genomes often uses gene panels with a series of strict variant filtering rules to identify likely disease-causing variants in monogenic disease genes [1–3]. Numerous gene curation initiatives exist [4] to evaluate gene-disease relationships, and lists of approved genes may be combined to create panels for comprehensive diagnostic testing of individuals presenting with appropriate clinical phenotypes. Panels can vary enormously in size - from several up to several thousand genes - depending on the heterogeneity of the clinical presentation of the disease and the evidence thresholds required for inclusion. The size and gene context of a panel affects the specificity and sensitivity of the test, but these standard test performance characteristics are rarely investigated. Although studies have shown that bigger gene panels aren’t necessarily better [5–7], nonetheless a focus on increasing diagnostic yields can result in a tendency towards ever larger panels.

We sought to investigate how gene panel size affects the background rate of candidate variants prioritised by applying standard rare disease variant filtering pipelines in a population setting, where the majority of variants will be false positives, i.e., not highly penetrant causes of monogenic disease. Using exome sequencing data from UK Biobank (UKB), which is known to have a strong ascertainment bias towards healthy adults [8,9], we tested five clinically curated gene panels that were developed to diagnose a range of heterogeneous monogenic diseases. We compared a subset of the results with data from the Deciphering Developmental Disorders (DDD) study, which includes families affected by severe undiagnosed developmental disorders [10,11].

## Methods

The Genotype-2-Phenotype plugin for the Variant Effect Predictor (G2P-VEP) [12] aims to identify likely disease-causing variants in a standard variant call format (VCF; https://samtools.github.io/hts-specs/) file based on a list of candidate genes. The plugin takes a list of input genes, along with their annotation as monoallelic or biallelic. It uses VEP [13] computed consequence of variants in an individual VCF file to identify non-synonymous variants within the input genes with gnomAD allele frequencies [14] less than 0.0001 for monoallelic genes or 0.005 for biallelic genes. Biallelic genes also require an individual to have two or more such variants within the gene so be identified as likely disease-causing. We used the G2P-VEP to identify rare non-synonymous variants in exome sequence data from 200,643 individuals in individuals in UKBB [15]. We limited our analysis to single nucleotide variants (SNVs) and small insertion/deletions on the autosomes. Ancestry was determined genetically by principal components analysis. Individuals were clustered using k-means clustering, and those clustering with individuals from the “EUR” group in 1000 Genomes were defined as European. We investigated five clinically curated panels of known monogenic disease-causing genes: developmental disorders (DD), Cancer, Cardiac, Skin, and Eye (downloaded from https://www.ebi.ac.uk/gene2phenotype, 26th June 2021; Cardiac panel, personal communication, 23rd July 2021; Supplementary Tables 1 to 5).

For each individual in UKB, we examined the number of variants in genes within each panel that were prioritised by the plugin. For each gene in the panel, we also examined the number of individuals who had prioritised variants. We estimated the expected number of individuals with variants identified within a given gene based on the length of the coding sequence of the canonical transcript of the gene. For monogenic genes, we fitted a simple linear regression line:

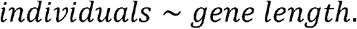

For biallelic genes, where two rare non-synonymous variants are required in each individual, we fitted the data to a quadratic model:

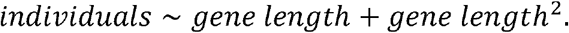

To assess the impact of overlapping phenotypes in common disease, of which a small subset might be caused by monogenic disease variants, we tested the difference between variants prioritised in cancer cases verses controls using the UKB cancer registry data. We then compared the distribution of variants identified within cases and controls separately within the Cancer gene panel.

Given the widespread use of pathogenicity predictors and clinical databases in diagnostic variant classification [16], we further assessed the impact of using REVEL scores [17] and ClinVar [18] variant classifications provided in the G2P-VEP plugin output files. Quintiles of REVEL score were calculated based on all variants prioritised by G2P-VEP within each gene panel separately. For monoallelic genes, variants were classified based on the highest REVEL score or ClinVar classification of all variants prioritised within each gene for each individual. For biallelic genes, variants were classified based on the second highest REVEL score or ClinVar classification of the two variants with the highest consequences within each gene for each individual.

To assess the diagnostic benefits of using the largest gene panel in a disease cohort, as well as the benefit of having haplotype data, we tested the number of variants prioritised using the DD gene panel in exome sequence data from 9859 proband-parent trios in the DDD study [10,11]. Ancestry was determined genetically by principal components as described previously [19]. For individuals with at least wo heterozygous variants prioritised in biallelic genes we examined parental genotype to determine allelic transmission. Individuals with at least one variant unambiguously inherited from each parent were classified as having variants in trans, i.e., true compound heterozygotes. Where all variants prioritised for the individual within the gene were unambiguously inherited from a single parent, they were classified as being in cis. Where individuals could not be classified by either of these rules, they were classified as unknown transmission.

## Results

The number of genes in each panel is shown in Table 1 along with the total length of the genes in each list. Notably, the largest panel includes over 7.3% of the exome. Figure 1 shows the number of variants prioritised in each individual in UKB compared to the number of genes in the gene panel. As anticipated, panels containing more genes resulted in more variants per individual being prioritised. The mean number of variants prioritised per individual ranged from 0.3 in the Cancer and Cardiac panels, to 3.5 in the DD panel (Table 1).

**Table 1:** Number of genes in each gene panel, plus mean and range of variants prioritised per person for each gene list.

| Panel | Number of Genes | Biallelic Genes | Monoallelic Genes | Total length (bp) | Percentage of Exome | Mean (minimum-maximum) number of rare variants prioritised in UKB | Genes with zero UKB individuals |
| --- | --- | --- | --- | --- | --- | --- | --- |
| Cancer | 91 | 37 | 62 | 238,608 | 0.4% | 0.3 (0-8) | 0 |
| Cardiac | 49 | 7 | 43 | 229,764 | 0.4% | 0.3 (0-8) | 0 |
| DD | 1708 | 1130 | 644 | 4,323,149 | 7.3% | 3.5 (0-37) | 57 |
| Eye | 536 | 408 | 178 | 1,343,235 | 2.3% | 1.3 (0-24) | 16 |
| Skin | 293 | 189 | 128 | 750,673 | 1.3% | 0.9 (0-14) | 11 |

**Figure 1:**
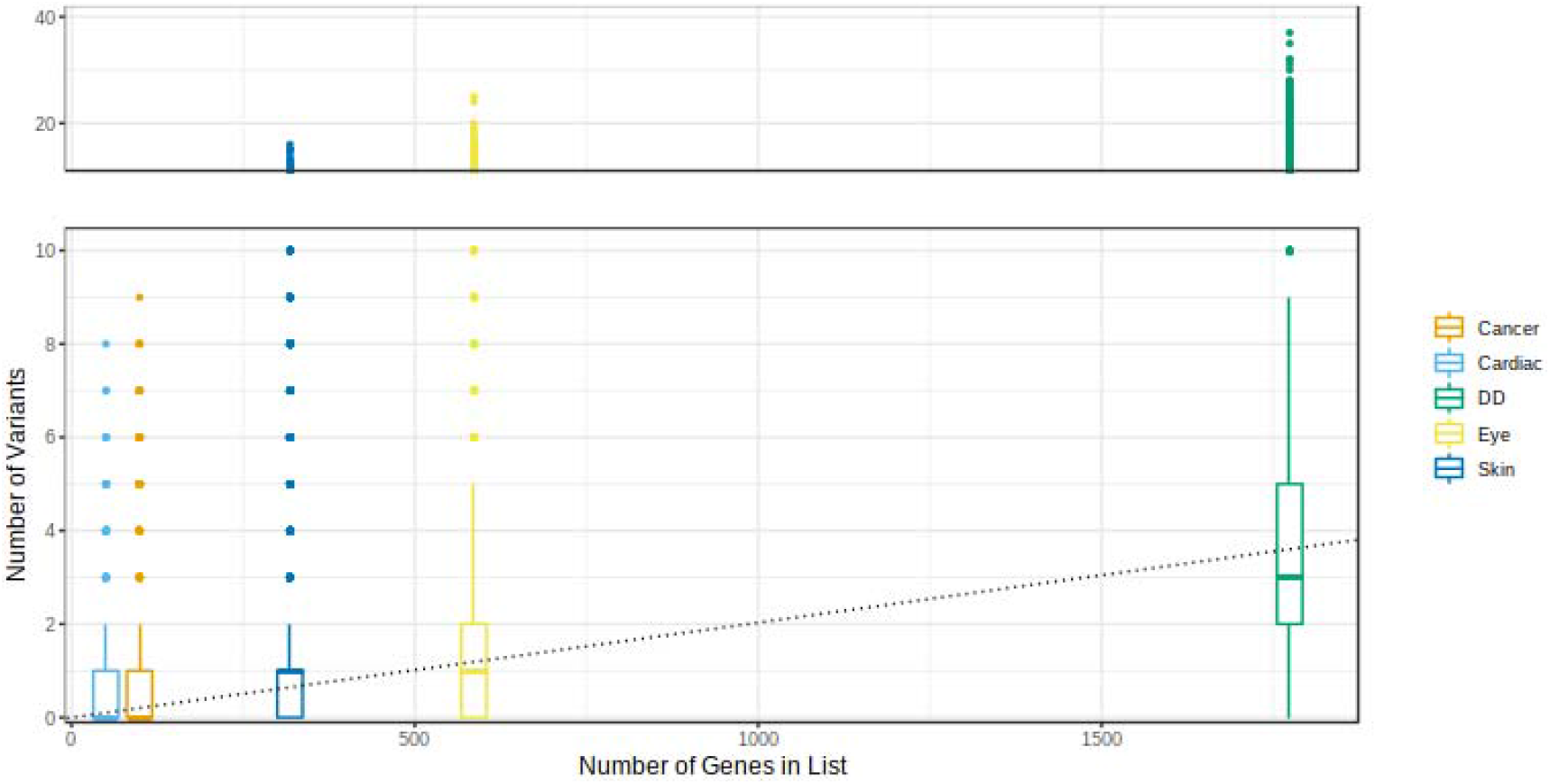
Number of rare non-synonymous variants prioritised by the VEP-G2P plugin in each person in UKB for each gene panel vs number of genes in panel.

Figure 2 shows histograms of the number of individuals who had variants prioritised within each gene for each panel. The distribution appears similar across all gene panels tested, suggesting that the majority of variants prioritised represent random noise rather than pathogenic variants. To examine the effect of overlapping common disease within the UKB we compared these distributions for the Cancer panel within cancer cases and controls separately (Supplementary Figure S1). We found no evidence for a difference in distribution between cases and controls (P=0.903), presumably due to the predominance of sporadic disease in this cohort.

**Figure 2:**
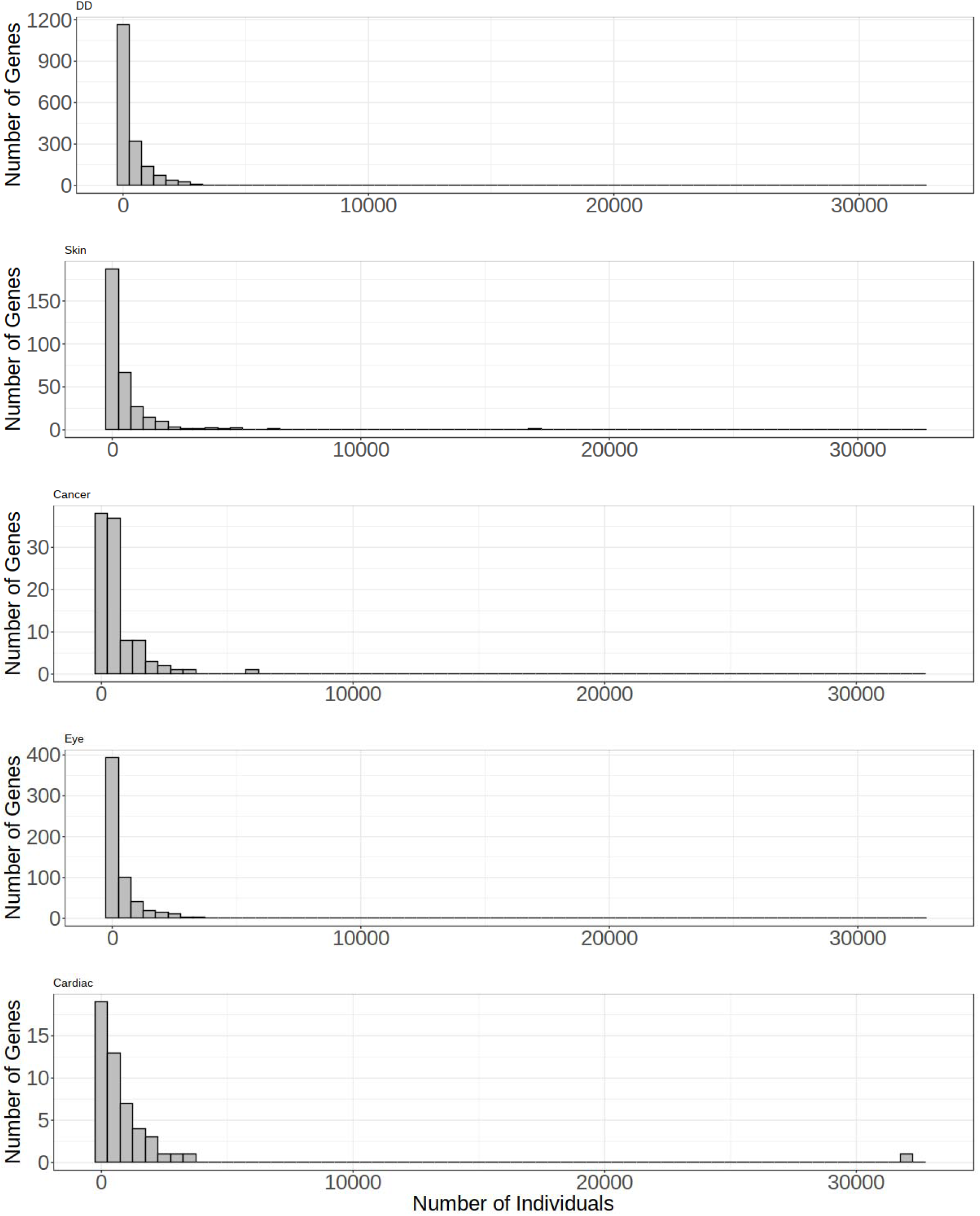
Histogram of number of individuals in UKB with variants within each gene in each panel. The width of each bin is 500 individuals.

For monoallelic disease genes across all panels, the number of variants prioritised in each gene increased linearly with the length of the canonical transcript of the gene (Figure 3 and Supplementary Figure S2); for each 1000 additional base pairs, there are an average of 295.8 additional variants prioritised. For biallelic disease genes, the number of variants scaled quadratically with the coding sequence length (Figure 3). For both monoallelic and biallelic genes there were notable outliers, which we defined as >3 SDs from the regression line, as well as well-known large genes that resulted in a disproportionately large number of prioritised variants (such as *NEB, SYNE1* and *TTN*; Table 2).

**Figure 3:**
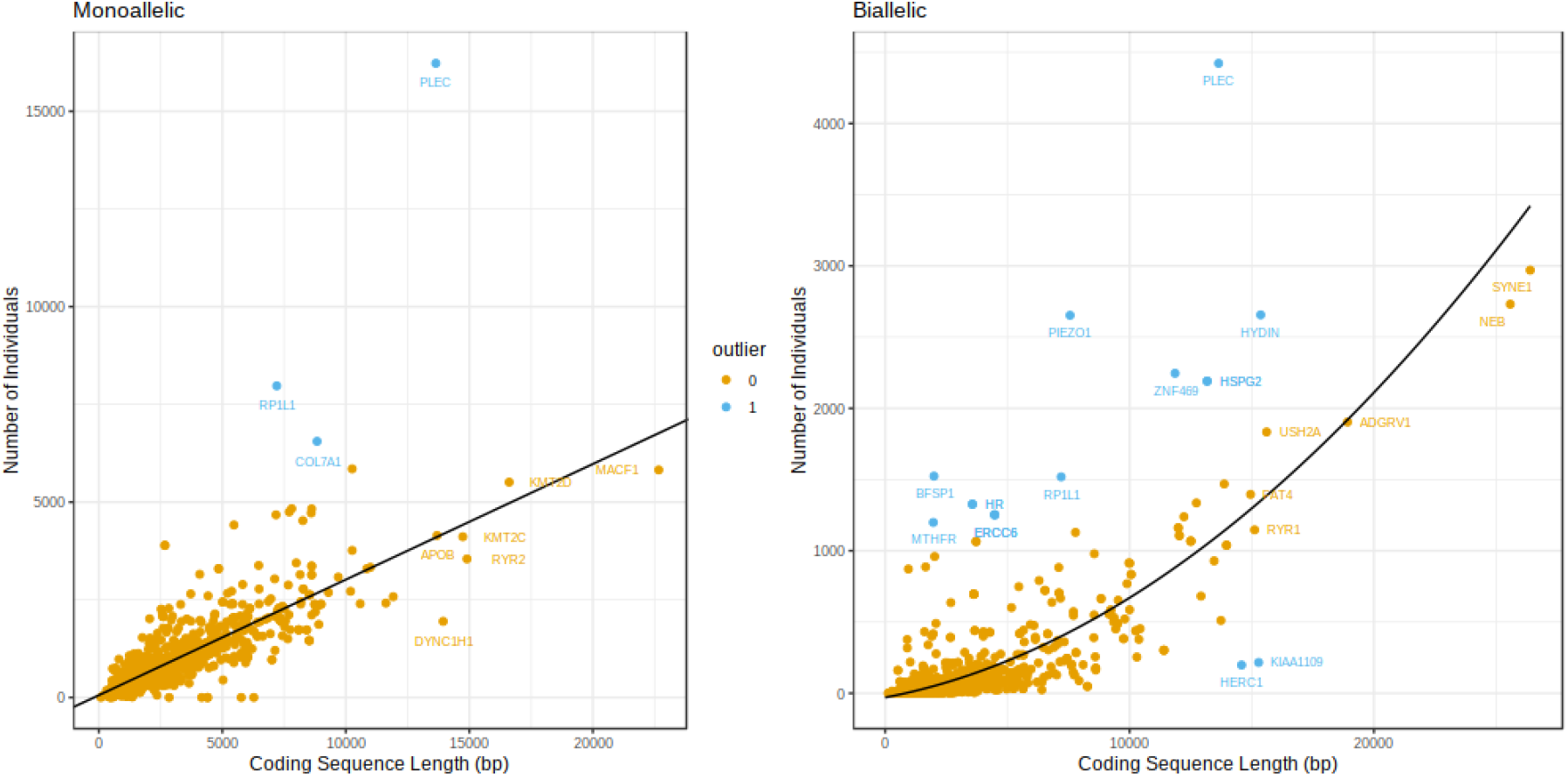
Coding sequence length of gene vs number of individuals with at least 1 variant (monoallelic) or 2 variants (biallelic) in the gene. Lines of best-fit are linear for monoallelic and quadratic for biallelic genes. Genes >3 SD from the regression line are highlighted in blue. The x-axis of the monoallelic genes has been truncated to increase resolution. The full figure can be found in Figure S2.

**Table 2:** Notable genes across all panels, highlighted either because they lie >3SDs outside of the trendline, or are very large (coding length of canonical transcript >20,000 bp) resulting in large numbers of variants being prioritised.

| Gene | Individuals | Length (bp) | Panel(s) | Type | Reason |
| --- | --- | --- | --- | --- | --- |
| *RP1L1* | 7974 | 7203 | Eye | Monoallelic | Outlier (high) |
| *PLEC* | 16227 | 13644 | Skin | Monoallelic | Outlier (high) |
| *COL7A1* | 6554 | 8835 | Skin | Monoallelic | Outlier (high) |
| *HYDIN* | 2656 | 15366 | DD | Biallelic | Outlier (high) |
| *HSPG2* | 2191 | 13176 | DD | Biallelic | Outlier (high) |
| *PIEZO1* | 2653 | 7566 | DD | Biallelic | Outlier (high) |
| *MTHFR* | 1199 | 1971 | DD | Biallelic | Outlier (high) |
| *ERCC6* | 1253 | 4482 | DD, Eye, Skin | Biallelic | Outlier (high) |
| *HR* | 1327 | 3570 | DDG, Skin | Biallelic | Outlier (high) |
| *HSPG2* | 2191 | 13176 | Eye | Biallelic | Outlier (high) |
| *ZNF469* | 2246 | 11862 | Eye | Biallelic | Outlier (high) |
| *RP1L1* | 1519 | 7203 | Eye | Biallelic | Outlier (high) |
| *BFSP1* | 1524 | 1998 | Eye | Biallelic | Outlier (high) |
| *PLEC* | 4422 | 13644 | Skin | Biallelic | Outlier (high) |
| *KIAA1109* | 216 | 15282 | DD | Biallelic | Outlier (low) |
| *HERC1* | 198 | 14586 | DD | Biallelic | Outlier (low) |
| *TTN* | 30308 | 107976 | Cardiac | Monoallelic | Gene size |
| *MACF1* | 5823 | 22668 | DD | Monoallelic | Gene size |
| *SYNE1* | 2970 | 26394 | DD | Biallelic | Gene size |
| *NEB* | 2731 | 25578 | DD | Biallelic | Gene size |

Comparing individuals of European ancestry to those of non-European ancestry within UK Biobank we found that individuals of non-European ancestry had a larger number of variants prioritised on average than individuals of European ancestry (5.4 and 3.3 respectively). The majority of the variants prioritised by G2P-VEP were missense variants with an average of 3.4 missense variants prioritised per gene, compared with just 0.1 predicted loss of function (LoF) variants (including stop gain, splice acceptor, splice donor and frameshift variants; Table 3). To investigate the potential for using additional evidence to reduce the number of prioritised missense variants, we separated the prioritised variants based on quantile of REVEL score or ClinVar pathogenicity classifications (Figure 4). There were substantially more variants prioritised with REVEL scores in the lowest quintile than in the highest quintile suggesting that the number of variants highlighted by the pipeline can be reduced by using REVEL or other appropriate pathogenicity predictors [20]. Examining the ClinVar classifications of the variants prioritised showed that more of the prioritised variants were classified in ClinVar as benign/likely benign than as pathogenic/likely pathogenic, highlighting the benefit of using clinical variant databases in variant prioritisation.

**Table 3:** Subsets of mean, median, minimum, and maximum variants prioritised in the DD panel per individual in the DDD cohort compared with UKB. (EUR = European, LoF = loss-of-function).

| Cohort | Mean | SD | Median | Minimum -<br>Maximum | N |
| --- | --- | --- | --- | --- | --- |
| UKB | 3.5 | 2.3 | 3 | (0 - 37) | 200643 |
| UKB EUR | 3.3 | 2.1 | 3 | (0 - 16) | 184477 |
| UKB non-EUR | 5.4 | 3.5 | 5 | (0 - 37) | 16155 |
| UKB LoF | 0.1 | 0.3 | 0 | (0 - 5) | 200643 |
| UKB missense | 3.4 | 2.3 | 3 | (0 - 36) | 200643 |
| UKB monoallelic genes | 2.6 | 1.7 | 2 | (0 - 21) | 200643 |
| UKB biallelic genes | 0.9 | 1.5 | 0 | (0 - 26) | 200643 |
| DDD parents (unaffected) | 4.9 | 3.7 | 4 | (0 - 163) | 17077 |
| DDD probands | 5.5 | 3.5 | 5 | (0 - 104) | 9859 |
| DDD probands EUR | 5.0 | 3.0 | 5 | (0 - 104) | 8522 |
| DDD probands non-EUR | 8.6 | 4.5 | 8 | (0 - 39) | 1337 |
| DDD probands LoF | 0.4 | 0.8 | 0 | (0 - 9) | 9859 |
| DDD probands missense | 4.5 | 3.2 | 4 | (0 - 102) | 9859 |
| DDD probands monoallelic genes | 3.5 | 2.2 | 3 | (0 - 78) | 9859 |
| DDD probands biallelic genes | 2.1 | 2.5 | 2 | (0 - 29) | 9859 |
| DDD (variants in cis in biallelic<br>genes) | 1.1 | 1.4 | 0 | (0 - 13) | 9859 |
| DDD (variants in trans in biallelic<br>genes) | 0.6 | 1.1 | 0 | (0 - 7) | 9859 |

**Figure 4:**
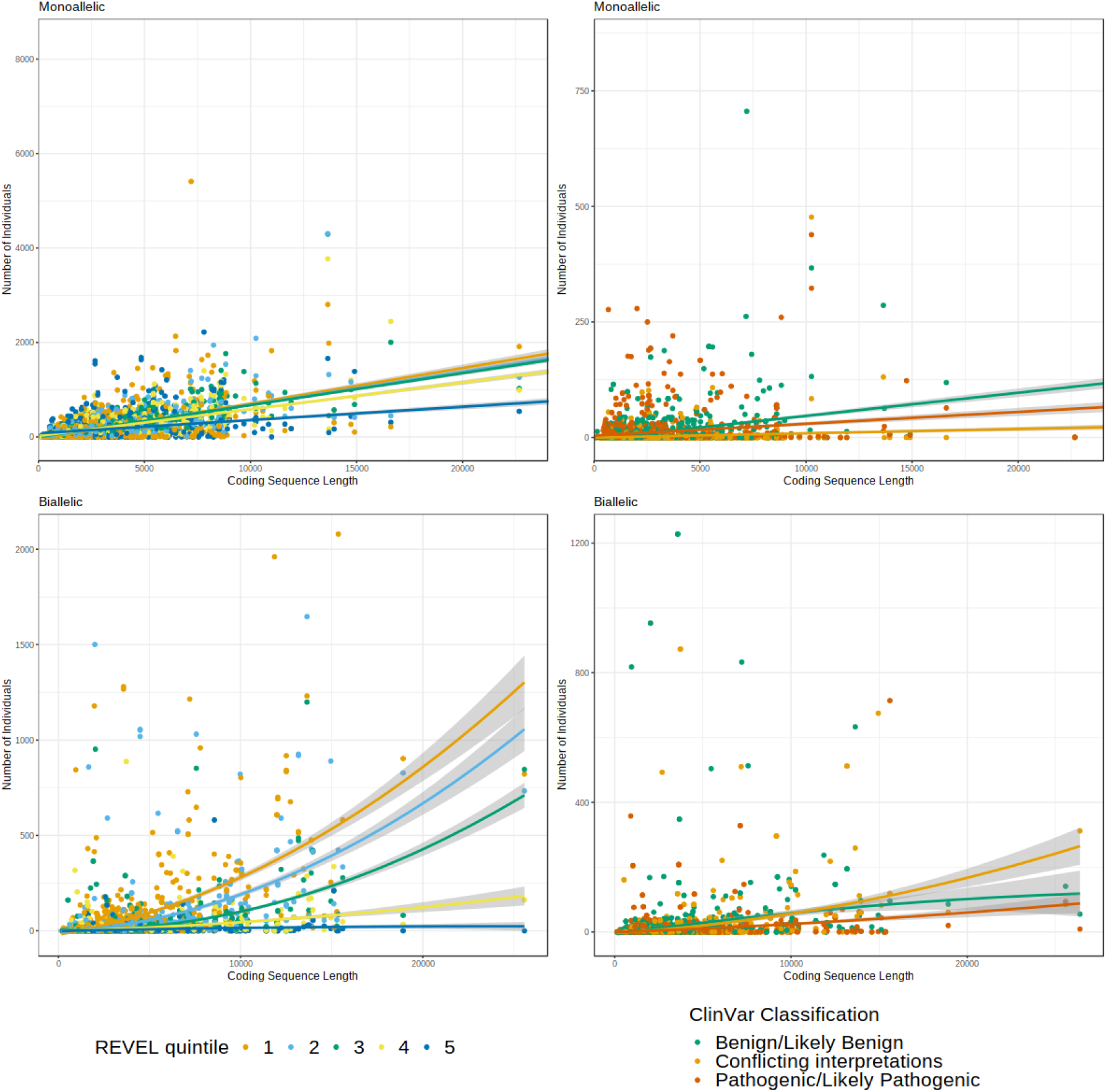
Scatter plot of length of gene against number of individuals with variants identified in that gene, split by REVEL quintile (left) or ClinVar classification (right) of the variant with the most severe consequence (Monoallelic, top) or the least severe consequence of the 2 highest consequence variants (Biallelic, bottom). The x-axis of the monoallelic genes has been truncated to increase resolution.

Using just the largest panel (DD), there were substantially more variants prioritised per individual for probands and parents in the DDD cohort compared to UKB (Table 3). This difference is likely to be due to the different study designs between UKB (a cross-sectional population cohort) and the DDD study (a disease cohort), resulting in opposite recruitment biases, though may also be due in part to technical reasons such as differences in the exome sequencing pipelines or reference genome builds between the two cohorts. Within the DDD cohort, probands had more variants on average (5.5) than the parents (4.9), the former likely reflecting the high rate of pathogenic de novo mutations in the cohort, and the latter likely reflecting incomplete penetrance in dominant conditions. In common with the UK Biobank, individuals with genetically determined non-European ancestry had more prioritised variants than those with European ancestry (Table 3). Using parental information to determine allelic transmission for biallelic genes in the DDD cohort (i.e., to separate individuals for whom prioritised variants within a gene all appeared in cis from those where at least two variants were in trans) we found that on average there were 0.3 genes per individual with variants in trans per individual on average, compared to 0.5 with variants in cis (Supplementary Figure S3).

## Discussion

The aim of this study was to estimate the diagnostic noise in panel-based genomic analysis by quantifying the normal background rate of rare non-synonymous variants in monogenic disease genes compared with the genomic footprint of the panel. We used G2P-VEP to estimate the number of potentially disease-causing variants prioritised in a clinically unselected population of individuals from the UK Biobank. We found that on average, for every 100 genes in a gene-panel, there were ∼0.2 variants prioritised in each individual, increasing linearly with the number of genes in the panel. This finding suggests that for larger gene panels (>500 genes), almost every individual tested will have at least one rare non-synonymous candidate diagnostic variant that is likely to be a false positive, i.e., variant that does not cause disease in that individual. Restricting missense variants to just ClinVar pathogenic/likely pathogenic or REVEL >0.7 (in keeping with variant interpretation guidelines from the UK Association of Clinical Genomics Scientists [21]) substantially reduces the number of background candidate variants, but still results in around ∼0.003 variants prioritised per individual per 100 genes in the panel. Therefore, even using these very conservative thresholds, we estimate that panels including all monogenic disease genes (∼5000 genes; the so-called ‘Mendeliome’ [22]) would produce a false positive result in around one in every six individuals tested.

Comparing the number of individuals with variants prioritised within each gene across all panels revealed that this number increased linearly with the length of the coding sequence for monoallelic genes, and quadratically for biallelic genes. We identified numerous genes which fell outside of these trends (Figure 3 and Table 2). Outlying genes included highly variable genes with more variants prioritised than expected based on their size (e.g., *PLEC*), and two highly constrained biallelic genes (*KIAA1109* and *HERC1*) with substantially fewer variants than expected. Genes with more identified individuals than expected warrant caution, even if identified in affected individuals, as they may be more likely to produce false-positive results. Conversely, biallelic variants in *KIAA1109* or *HERC1* may be more likely to be true diagnoses in affected individuals as they are found at a lower rate in the general population.

To the best of our knowledge, this is the first study that attempts to quantify the normal background rates of candidate disease-causing variants in monogenic disease gene panels. We believe our findings are extremely relevant to clinical and research diagnostic variant filtering following exome or genome sequencing for heterogenous monogenic diseases. Although the exact details of variant filtering for rare disease diagnosis differ between laboratories, there are some standard practices that are universally accepted and shared by most pipelines as well as the G2P-VEP plugin. Firstly, individual variants must be rare, with allele frequency thresholds that depend upon mechanism of disease; the thresholds used in the G2P-VEP plugin (<0.0001 for monoallelic and <0.005 in biallelic genes) are conservative as they were developed for use in developmental disorders, where causal variants are likely to be depleted in the population. Secondly, variants must have an interpretable functional effect, which in practice usually means that non-synonymous coding variants are selected for inclusion. Thirdly, although a single variant alone is sufficient to cause monoallelic disease, two variants in trans in the same person are required to cause biallelic disease; in the absence of inheritance or long-read sequence data, the phase of these candidate biallelic variants is usually unknown. These rules are all requirements of the G2P-VEP plugin. In addition, many pipelines will also use pathogenicity prediction scores (such as REVEL) or presence in clinical databases (such as ClinVar) as a further filtering step, which we have also replicated in this study. Strengths of our analyses include the size of the UK Biobank, and the population-based nature of the cohort. The majority of the participants are healthy and unaffected by the diseases represented by the panels tested here. Sensitivity analysis comparing cancer cases to controls suggest that the presence of undiagnosed cases in the cohort is unlikely to have a large effect on the results presented here, although the presence of some relevant monogenic disease cases in the cohort [23,24] remains a limitation of our approach.

The growing interest in newborn sequencing and differences in the genes included in panels used [25] means that estimating the background distribution of likely benign variants identified by panel-based genomic analysis is essential both for appropriate gene selection and interpretation of the results. While panel-based analysis is an efficient way of identifying possibly pathogenic variants in known monogenic genes, many of the variants prioritised by such approaches will not be disease causing and as the genomic footprint of the panel increases, so too will the number of prioritised variants. As with any test, there is a trade-off between sensitivity and specificity; in the case of rare diseases, including more genes on a panel, or using less strict variant filtering rules, will result in more diagnoses (higher sensitivity) but also more false positives (lower specificity). Due to the bias towards Europeans in existing cohorts, these issues are more pronounced for individuals of non-European origin (in whom a greater number of variants will be prioritised due to inaccurate allele frequency annotations), highlighting the need for greater ethnic diversity amongst sequenced populations to improve diagnostic accuracy [26].

## Conclusions

Although large gene panels may be the best strategy to maximize diagnostic yield in genetically heterogeneous diseases, they will frequently prioritise false positive candidate variants potentially requiring additional clinical follow-up. Our findings highlight the importance of careful phenotypic selection of individuals for panel testing to improve clinical specificity. Our findings also support the need for sequence data from more ethnically diverse groups to improve variant filtering in non-Europeans. Overall, we suggest that extreme caution should be applied when interpreting potentially pathogenic variants found incidentally.

## Data Availability

All data produced in the present work are contained in the manuscript

## Ethics, consent and permissions

The UK Biobank has approval from the North West Multi-centre Research Ethics Committee (21/NW/0157) as a Research Tissue Bank approval. The DDD study has UK Research Ethics Committee approval (10/H0305/83, granted by Cambridge South REC, and GEN/284/12 granted by the Republic of Ireland REC). All individuals included in this study gave appropriate consent.

## Acknowledgements

The authors wish to thank Andrew R Wood for assistance with the UKB exome sequencing data, James Ware for providing the Cardiac gene panel, and Helen Firth and David FitzPatrick for helpful suggestions. This work was supported by the MRC [MR/T00200X/1] and the Wellcome Trust [200990/A/16/Z] and was conducted using the UK Biobank Resource under Application number 49847. The DDD study presents independent research commissioned by the Health Innovation Challenge Fund [grant number HICF-1009-003] a parallel funding partnership between the Wellcome Trust and the Department of Health, and the Wellcome Trust Sanger Institute [grant no. WT098051]. See Nature 2015:519”223-8 or www.ddduk.org/access.html for full acknowledgement. The authors would like to acknowledge the use of the University of Exeter High-Performance Computing (HPC) facility in carrying out this work. The views expressed in this work are those of the authors and not necessarily those of the funders. For the purpose of open access, the author has applied a CC BY public copyright licence to any Author Accepted Manuscript version arising from this submission.

## Conflicts of Interest

The authors have no conflict of interest

## Supplemental Figures

**Supplemental Figure 1:**
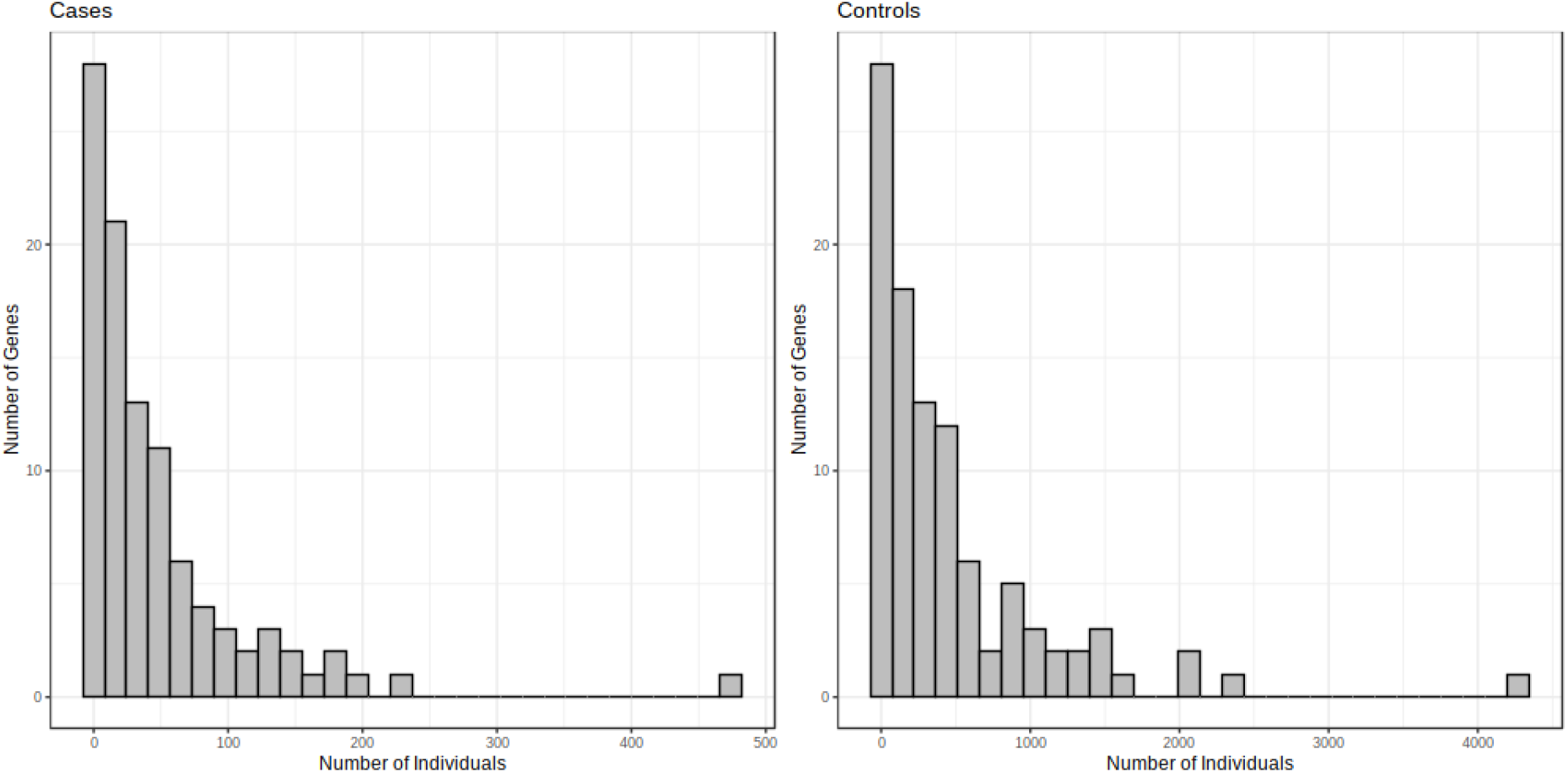
Histograms of the number of individuals with variants within each gene in the Cancer panel, split by cases and controls.

**Supplemental Figure 2:**
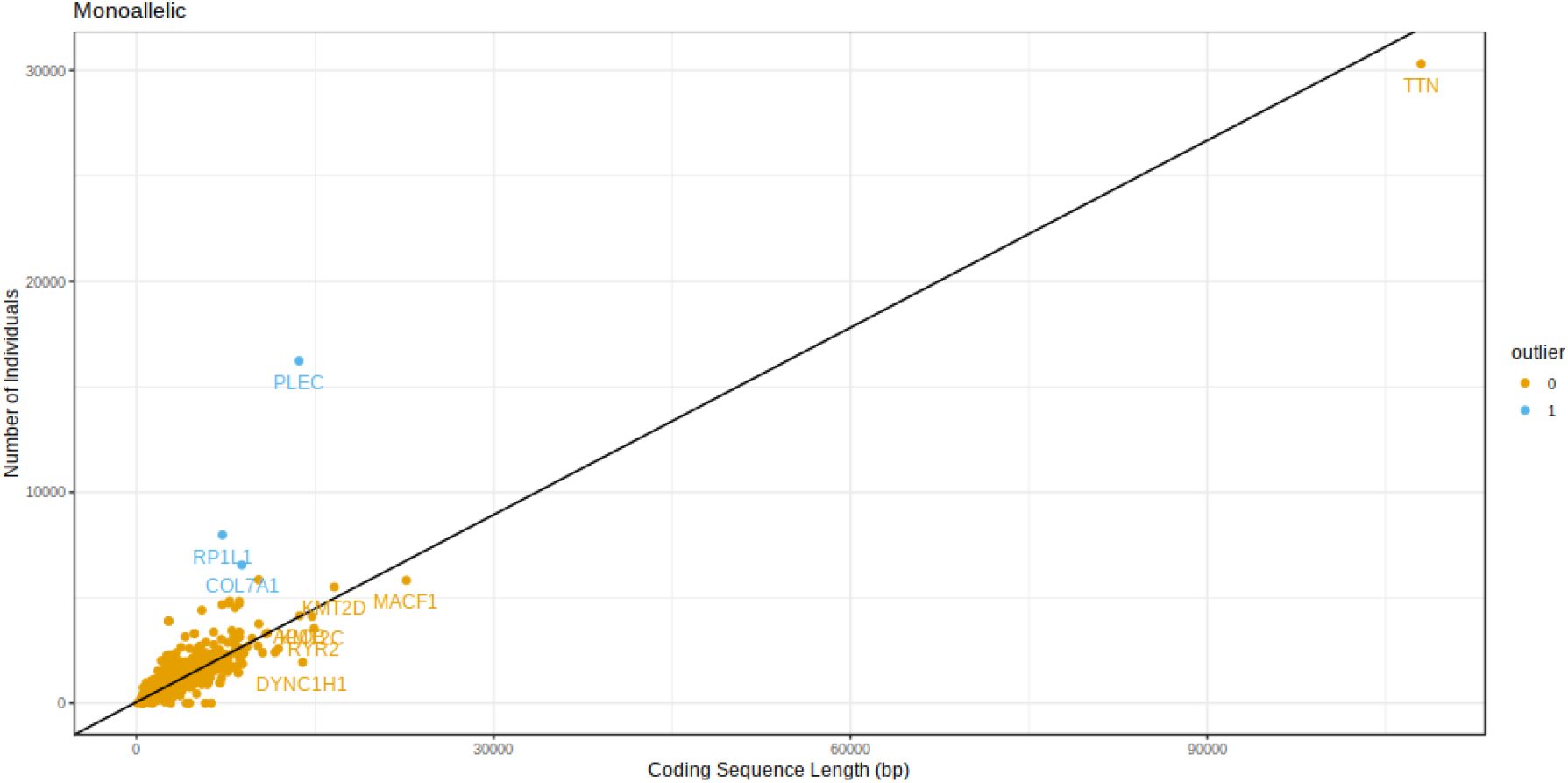
Coding sequence length of gene vs number of individuals with at least 1 variant in the monoallelic genes. Genes further than 3 SD from the regression line are highlighted in blue.

**Supplemental Figure 3:**
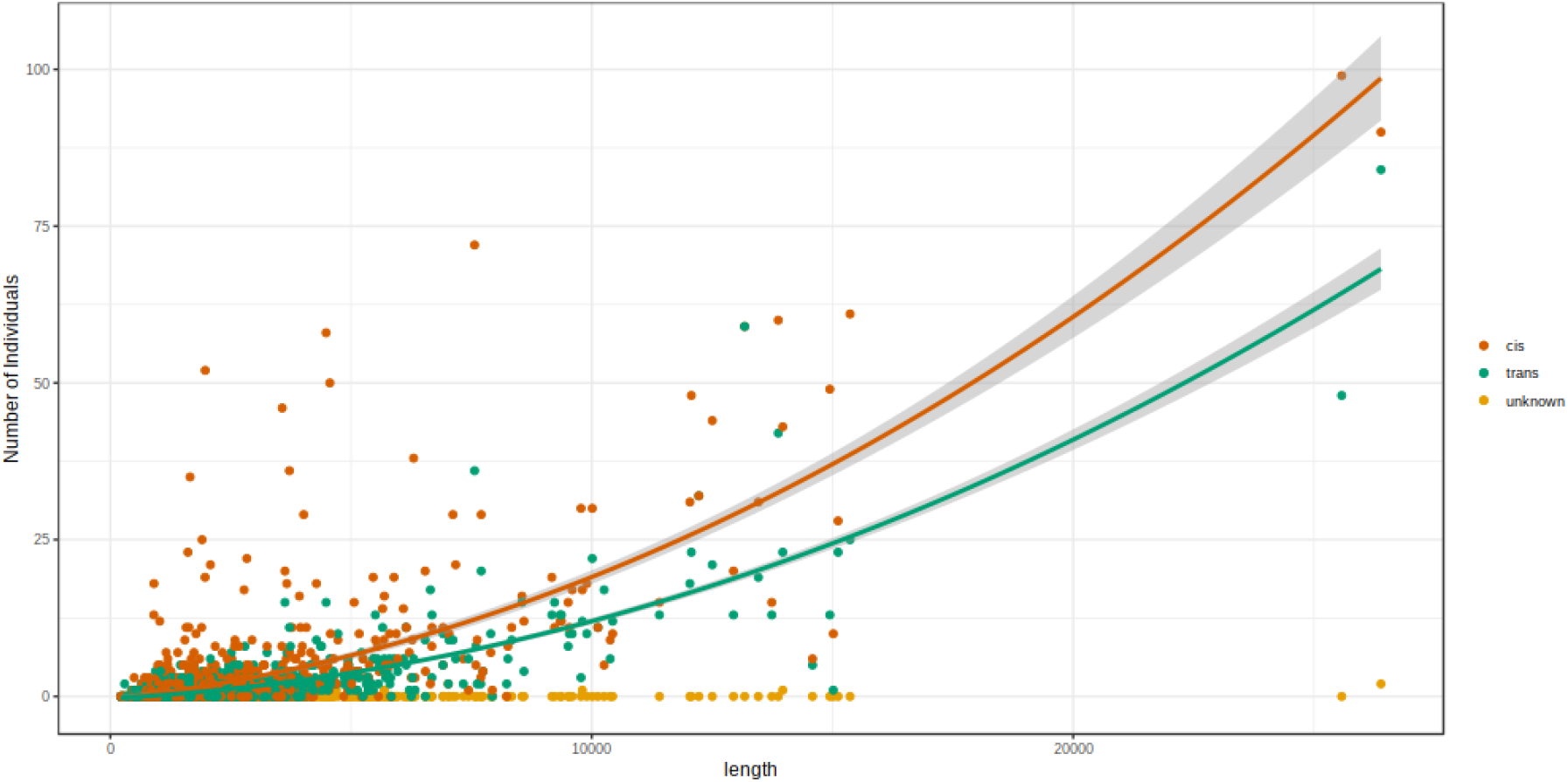
Scatter plot of length of gene against number of variants identified in the gene for biallelic genes for individuals in the DDD cohort, split by whether they appear in cis, trans, or undetermined based on parental genotype.

## Notes

### Competing Interest Statement

The authors have declared no competing interest.

